# The Role of Electrode Placement in STN-DBS for Improving Gait in Parkinson’s Disease

**DOI:** 10.1101/2023.12.15.23299998

**Authors:** Zhongke Mei, Anna-Sophie Hofer, Christian Baumann, Mechtild Uhl, Navrag Singh, William R. Taylor, Lennart Stieglitz, Deepak Ravi

## Abstract

**Background:** Deep brain stimulation (DBS) targeting the subthalamic nucleus (STN) is a widely adopted therapy for alleviating motor symptoms in Parkinson’s Disease (PD). While electrode placement has been proposed as a key factor influencing motor outcomes, the specific relationship between electrode location and therapeutic effects on gait performance remain unclear. This study investigates the role of electrode placement in STN-DBS for improving gait in patients with PD (PwPD).

**Methods:** We conducted an observational study to evaluate overground gait performance in 49 PwPD who underwent bilateral STN-DBS surgery. Gait assessments were performed both prior to treatment initiation and six months post-implantation. We analysed changes in the mean values (mean set) and coefficient of variability (variability set) of ten commonly used spatio-temporal gait parameters, including stride length and walking speed. Additionally, we explored the association between gait outcomes and the spatial location of active contacts of the DBS electrodes.

**Results:** Our findings indicate that DBS treatment resulted in a significant reduction in stride time, stance time, swing time, and step time, in addition to an increase in the variability of double limb stance time, stride time, stance time, and step time. Furthermore, we observed that the location of the active contacts was associated with alterations in mean step length, stride length, and walking speed, as well as changes to cadence, stride time, stance time, and step width. We identified the postero-superior region of the STN as the most effective region for improving the mean set, whereas the antero-superior region of the STN emerged as the most effective region for improving the variability set.

**Conclusions:** This study provides empirical evidence on how STN-DBS, together with the spatial location of active lead contacts, impacts both the mean and variability of spatio-temporal gait parameters in PwPD. Importantly, our results highlight specific spatial targets within the STN that may optimize DBS implantation strategies to address patient-specific gait symptoms.

## 1. Introduction

Parkinson’s disease (PD) is the second-most common neurodegenerative disorder that affects 2-3% of the population ≥65 years of age[1]. The pathogenesis of PD involves neuronal loss in specific areas of the substantia nigra and widespread intracellular protein accumulation, leading to various pathological motor and non-motor symptoms, including significant gait impairments[2]. Deep brain stimulation targeting the subthalamic nucleus (STN-DBS) has been established as an effective treatment for managing motor symptoms in PD. Several studies have demonstrated that STN-DBS improves dopamine-responsive motor symptoms, as reflected in reduction in scores on the motor part (III) of the Unified Parkinson Disease Rating Scale (UPDRS), particularly akinesia, rigidity, and tremor[3–7]. However, the UPDRS Part III provides a limited and oversimplified assessment of gait symptoms. As a result, relying solely on these scores for surgical planning may contribute to variability in treatment outcomes for patients with PD (PwPD)[8, 9].

Despite the well-established efficacy of STN-DBS in alleviating motor symptoms such as tremor and bradykinesia, its impact on gait performance remains controversial[9–11]. Existing studies report mixed findings: while some demonstrate improvements in spatial aspects of gait such as increased step length and walking speed [12, 13], others suggest that STN-DBS may exacerbate temporal gait variability, potentially compromising walking stability[9] or slow down patient’s movement, suggesting a potential mechanism for gait deterioration under certain stimulation conditions [14]. Recent evidence further indicates that STN-DBS may influence gait variability through distinct mechanisms, with some studies showing a reduction in short-term variability of lower limb parameters, possibly medicated by non-dopaminergic pathways[15]. Consequently, the overall effects of STN-DBS on gait performance in PwPD remain unclear.

With advancements in imaging techniques and software for lead localization and reconstruction[16], the significance of active contact localization in determining therapeutic outcomes has become increasingly evident. Several studies suggest that the interface between the supero-dorso-lateral STN, zona incerta, and Forel’s fields plays a key role in mediating the motor effects of STN stimulation [17–20]. Importantly, recent evidence has demonstrated a correlation between gait improvement and the location of active contact within the STN of PwPD[21]. Furthermore, meta-analyses have identified distinct ‘sweet spots’ associated with improvements in specific motor symptoms. For example, stimulation at the supero-dorso-medial border of the STN is linked to improvements in rigidity and bradykinesia, the dorsolateral region targets tremor and rigidity, and stimulation at the ventral boundaries has been associated with gait improvements[22–25]. Based on this evidence, we hypothesize that stimulation location may also account for the variability in gait outcomes observed with STN-DBS. However, a comprehensive investigation into these spatial relationships and their specific impact on gait performance is still lacking.

In this study, we aim to comprehensively investigate the impact of STN-DBS and stimulation location on gait performance in PwPD. Specifically, we examine spatio-temporal gait parameters before (patients under best medication state) and six months after surgery (patients under optimal combined treatment state). We analyse changes in both the mean values and variability of key gait parameters to capture the full spectrum of gait performance.

Recognizing the significant role that electrode placement may play in these outcomes, we further assess the relationship between treatment-induced changes and the spatial localization of active stimulation contacts within the STN. Through this comprehensive investigation, we aim to identify the most effective stimulation regions for maximizing improvements in gait performance, thereby paving the way for more personalized DBS surgical strategies. Ultimately, the findings from this study are expected to contribute valuable insights into how spatial variability in electrode placement influences clinical outcomes, supporting the development of precision-guided neuromodulation treatments tailored to the specific gait deficits of PwPD.

## 2. Methods

### 2.1 Study Participants

A convenience sample of 49 PwPD (excluding 9 dropouts), who were scheduled for STN-DBS treatment, was recruited at the University Hospital of Zurich in the years 2016 - 2021. This single-center study was carried out with the expertise of an experienced neurosurgical team using a standardized protocol, thereby ensuring consistency in surgical technique and postoperative care across all procedures. The sample consisted of 41 male and 8 female patients, with a mean age of 59.7±10.4 years and a confirmed diagnosis of Parkinson’s disease with a disease course of 8.9±4.5 years. The study was approved (approval no: 201500141) by the Zurich Cantonal Ethics Commission and carried out in accordance with the Declaration of Helsinki, the guidelines of Good Clinical Practice, and the Swiss regulatory authority’s requirements. All participants provided written, informed consent prior to study involvement. The detailed surgical protocol and postoperative management of patients are provided in Supplementary Methods 1. Stimulation parameters are also presented in Supplementary Table 1.

### 2.2 Image acquisition and localization of active contact

Reconstruction of each electrode’s active lead contact localization within the left and right STN was performed using the Brainlab (www.brainlab.com) stereotactic planning software. Each patient’s native brain computed tomography (CT) scan and T2-weighted magnetic resonance imaging (MRI) scan (3D vista sequence) were automatically fused, including manual correction if required. Afterwards, automated anatomical mapping was performed, with accuracy of relevant anatomical structure mapping double-checked manually. Next, anterior-commissure to posterior-commissure (AC-PC) lines were inserted based on the patient’s MRI using the lead localization function, and electrodes were automatically detected based on the patient’s CT scans. For all detected electrodes, properties were set to the Medtronic 3389-lead type.

To address the substantial variability in STN size and shape across patients, we normalized the STN using a voxel-based approach. Specifically, the area in and around STN was divided into 5×5×7(=175) voxels, where the spatial localization of the active stimulation contacts relative to the STN were localized within one voxel of the 5×5×7-field grid instead of general stereotactic coordinates. This normalization allowed us to precisely localize the spatial positions of the active stimulation contacts within a consistent, patient-independent coordinate system, rather than relying solely on general stereotactic coordinates that may be affected by individual anatomical differences. The plane containing each electrode’s active contact centre of cathode stimulation (left electrode, contact 0-3 from electrode tip upwards, e.g., documented as 2-C+; right electrode, contact 8-11 from electrode tip upwards, e.g., documented as 9-C+) was selected in the x-direction (medio-lateral), y-direction (anterior-posterior) and z-direction (superior-inferior). In case two contacts were used for cathode stimulation, e.g., 2-3-C+, the plane centred in the middle between e.g., contact 2 and contact 3, was used for scoring.

*Active contact position in the X-direction was scored as follows:*

0 = contact completely outside of the STN medially; 1 = contact partially in the STN medially; 2 = contact in the central part of STN; 3 = contact partially in the STN laterally; 4 = contact completely outside of STN laterally.

*Active contact position in the Y-direction was scored as follows:*

0 = contact completely outside of STN anteriorly; 1 = contact partially in the STN anteriorly; 2 = contact in the anterior third of STN; 3 = contact in the central part of STN; 4 = contact in the posterior third of STN; 5 = contact partially in the STN posteriorly; 6 = contact completely outside of the STN posteriorly.

*Active contact position in the Z-direction was scored as follows:*

0 = contact completely outside of STN superiorly; 1 = contact partially in the STN superiorly; 2 = contact in the central part of STN; 3 = contact partially in the STN inferiorly; 4 = contact completely outside of STN inferiorly.

An illustrative example of scoring is shown in *Figure 1*.

**Figure 1.**
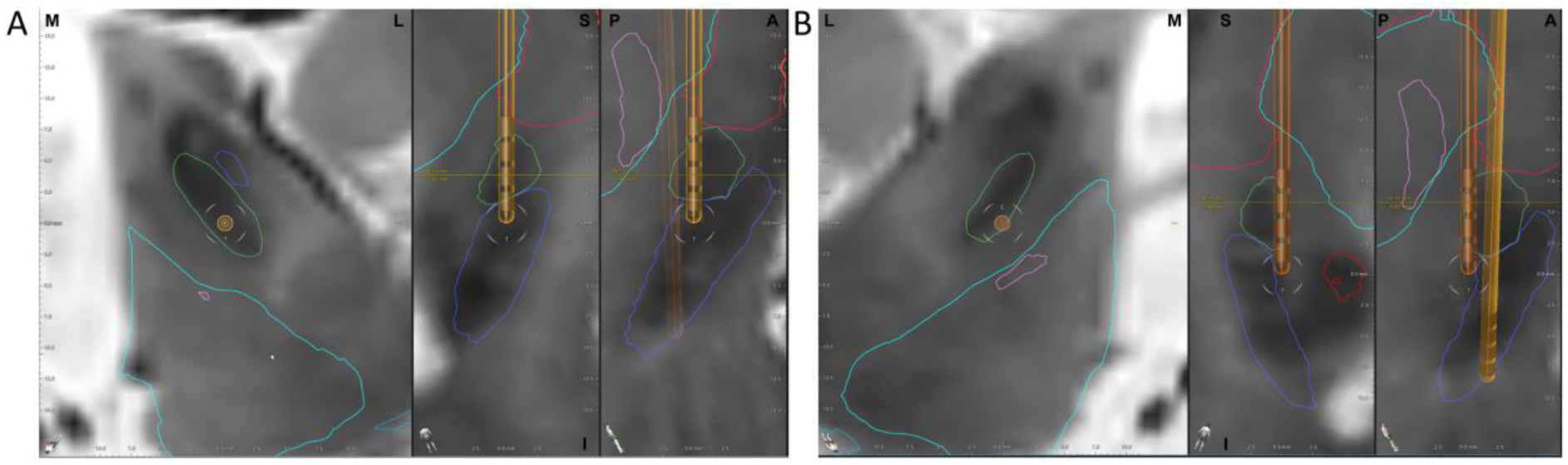
Exemplary location of active contacts of STN-DBS electrodes implanted into the (A) right hemisphere and (B) left hemisphere, where the light green lines indicate the boarders of the STN region. (A) Scoring of right-sided electrode with active contact 9 (9-C+, highlighted by horizontal line illustrating plane of analysis): medio-lateral score 2, superior-inferior score 2, anterior-posterior score 4. (B) Scoring of left-sided electrode with active contact 2 (2-C+, highlighted by horizontal line illustrating plane of analysis): medio-lateral score 1, superior-inferior score 2, anterior-posterior score 5.

### 2.3 Quantification of gait performance

Gait kinematics were assessed in 49 PwPD at two distinct treatment states. At baseline (Pre-DBS), patients were evaluated under their best medication state (ON medication) prior to DBS electrode implantation. Approximately six months following surgery, patients were re-assessed in their optimal combined treatment state (Post-DBS), with adjusted medication and active stimulation (ON medication and ON stimulation).

All participants walked barefoot for a continuous period of 10 minutes at a self-selected speed and without any assistance. Each participant was instructed to navigate an “8”-shaped path around two signs positioned 10 meters apart, with the aim of capturing consecutive gait cycles during overground walking in the laboratory setting[26]. Locomotion was recorded using a three-dimensional motion capture camera system (consisting of 10 cameras; 61 markers; sampling rate of 100 Hz; Vicon Nexus, version 2.3/2.8.2, Oxford Metrics, United Kingdom). Prior to recording, all cameras were calibrated with an error threshold set at 0.5 mm. Commonly used spatio-temporal parameters were calculated in MATLAB (version R2022a, The MathWorks Inc., Natick) by selecting the trajectories of the markers mounted to the participant’s heel (calcaneus), toe (metatarsal head III), and sacrum (midpoint between left and right posterior superior iliac spines).

In total, 6 temporal parameters (cadence, in steps/min; double limb support time [DLST], stride time, stance time, swing time, and step time, in s), 3 spatial parameters (step length, stride length, and step width, in cm), as well as walking speed (cm/s) were selected to represent the gait performance (Supplementary Methods 2). Outliers were identified and removed at the individual patient and parameter level. Specifically, for each patient and parameter, any step exhibiting values beyond ±4 median absolute deviations (MAD) from the median of that patient’s data was classified as an outlier and excluded from further analysis. The mean values and coefficients of variation (defined as standard deviation/mean x 100) of each parameter were then calculated to analyze changes between pre- and post-DBS surgery conditions.

### 2.4 Statistical analysis

To investigate the effects of DBS surgery on gait parameters, paired t-tests comparing Pre-DBS and Post-DBS datasets were conducted for both mean values and coefficients of variation, with significance level set at α=0.05. Gait parameters were analysed individually to assess whether the stimulation location had a parameter-specific effect. Therefore, no correction for multiple comparisons was applied in the statistical analysis. In order to provide a measure of precision and confidence about our estimates that is not sensitive to the distribution of the observations or underlying populations, effect sizes were calculated using 5000 times bootstrap resampling, along with the 95% confidence intervals[27]. The effect size was calculated using Hedges’ g and interpreted as a small effect if 0.2<g<0.5; a medium effect if 0.5≤g≤0.8; and a large effect if g>0.8[28, 29].

Next, the active contact locations were divided into three independent variables: the x-axis (from medial to lateral), y-axis (from anterior to posterior), and z-axis (from superior to inferior). For each gait parameter, differences between Post-DBS and Pre-DBS conditions were calculated to quantify the effect of DBS surgery. Subsequently, Kruskal-Wallis tests were conducted separately for each spatial axis (x, y, and z), using these parameter differences as dependent variables and voxel locations along each axis as independent factors (see Figure 3). Patients were grouped based on voxel positions corresponding to their active contacts. The significance level was set at α = 0.05. This analysis aimed to assess whether DBS-related improvements in gait parameters differed significantly according to stimulation location.

### 2.5 Most effective region investigation

Based on the Kruskal-Wallis test results, responsive parameters (those sensitive to contact location and exhibiting significant changes) were categorized into two sets: reduced mean and reduced variability. The average tendencies along the x-, y-, and z-axes were then calculated for each group. Subsequently, the most effective regions were visually defined based on these average tendencies. In case of the mean set, the most effective region was defined as the location where the maximum increase in spatial parameters occurred, indicating reduced bradykinesia and enhanced walking stability as individuals enhance their movement amplitude[8, 30, 31]. For the variability set, the most effective region was determined as the location where the maximum decrease (or minimal increase) in temporal parameters occurred, representing an increase (or limited decline) in walking stability for the patients[32]. Finally, t-tests were conducted for each selected parameter to determine whether there was a significant difference between patients with stimulation locations in the most effective region and those without, with a significance level of α=0.05. Statistical analyses were performed using MATLAB (version R2022a, MathWorks Inc.) and R (version 4.3.1, www.r-project.org).

## 3. Results

### 3.1 Patient demographics

The demographic characteristics, including the mean and standard deviation of age, sex distribution, weight, height, age at time of disease onset, and disease duration and severity (Hoehn and Yahr scale) of PwPD prior to treatment initiation and six months post-implantation, are shown in Table 1. Notably, none of the participants exhibited freezing of gait during the objective gait assessment, although 3 participants before the surgery and 5 participants after the surgery reported mild freezing of gait as indicated by their UPDRS scores.

**Table 1.** Baseline and Longitudinal Demographic Characteristics.

| Characteristics | PwPD Pre-DBS (N = 49) | PwPD Post-DBS (N = 49) |
| --- | --- | --- |
| Age (years) | 59.7 (10.4) | - |
| Male/Female (No.) | 41/8 | - |
| Weight (kg) | 76.8 (12.6) | 78.8 (11.9) |
| Height (cm) | 176.1 (6.8) | 175.7 (7.0) |
| Age at disease onset (years) | 50.8 (9.7) | - |
| Disease duration (years) | 8.9 (4.5) | - |
| H&Y stage | 2 (1–3) | - |
| MDS-UPDRS I | 9.76 (5.1) | 7.0 (3.3) |
| MDS-UPDRS II | 12.76 (4.3) | 8.0 (6.4) |
| MDS-UPDRS III | 18.9 (6.9) | 17.0 (7.9) |
| MDS-UPDRS IV | 7.62 (4.1) | 0.4 (2.6) |
| LEDD [mg] | 1060 (473.5) | 347.8 (295.9) |

Values are expressed as numbers of participants or mean (standard deviation). Abbreviations: PwPD, patients with Parkinson's disease; DBS, deep brain stimulation; H&Y, Hoehn and Yahr.

### 3.2 STN-DBS treatment effects on gait performance

The comparison of mean and variability of 10 gait parameters between groups revealed significant differences between Pre- and Post-DBS surgery (Figure 2).

**Figure 2.**
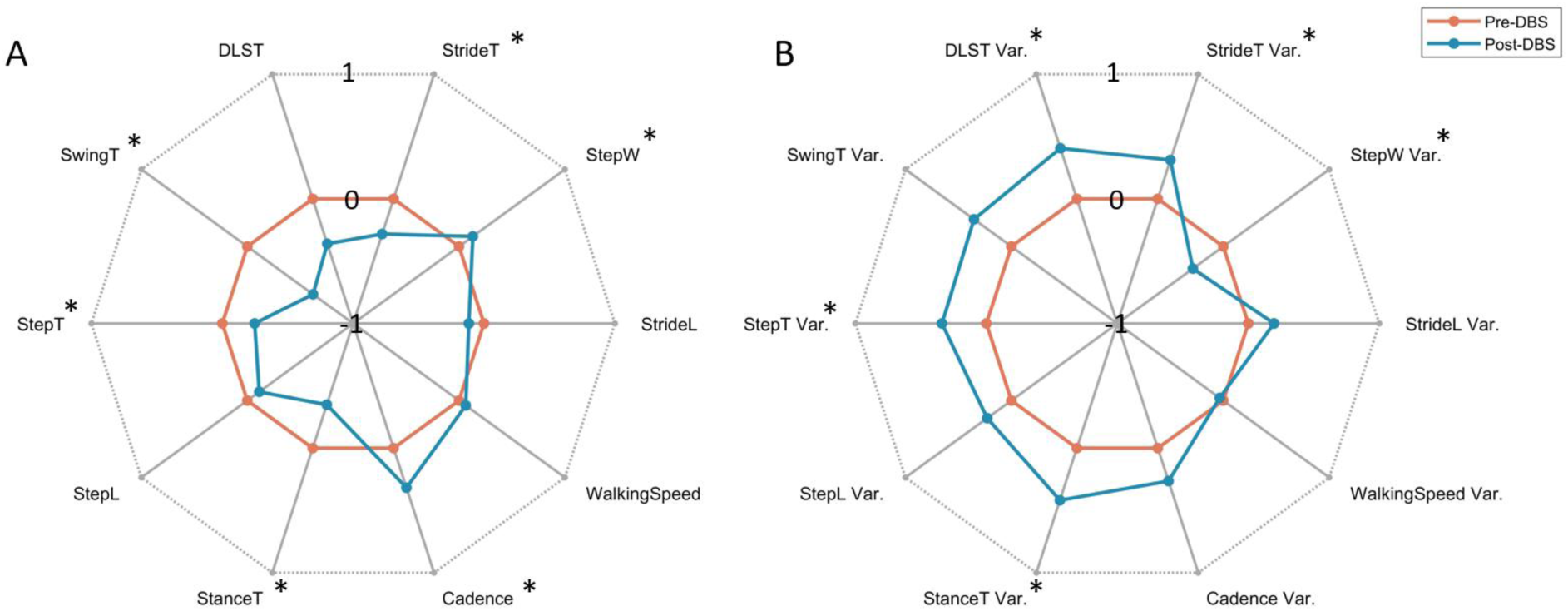
The radar plot illustrates (A) mean and (B) variability of 10 gait characteristics for PwPD before (Pre-DBS, red line) and 6 months after (Post-DBS, blue line) DBS surgery. The radial scale is based on ±1 times the standard deviation of the control group around the Z-score for each parameter, which was calculated based on the mean and standard deviation of the Pre-DBS group. For example, the mean cadence of PwPD before surgery is slightly smaller than that of PwPD after surgery. Asterisks represent significant differences between the Pre-DBS and Post-DBS groups.

Following DBS treatment, significant increases were observed in cadence and step width among the mean gait parameters, while stride time, stance time, swing time, and step time significantly decreased *(Table 2)*. Stride length and step length showed slight, but non-significant reductions. Walking speed and DLST did not differ between Pre-DBS and Post-DBS assessments.

**Table 2.** The means (SD) of gait parameters for Pre-DBS and Post-DBS group are depicted, accompanied by effect sizes with confidence intervals and p-values. Statistically significant differences are denoted with bolded numbers.

| Mean Gait Parameters | Pre-DBS | Post-DBS | p-value (pre-post) | Effect Size (Hedges' g, pre-post) | Effect Size Confidence Intervals |
| --- | --- | --- | --- | --- | --- |
| Cadence (steps/min) | 113.5 (7.7) | 115.8 (6.8) | **<0.001** | 0.318 | (0.090, 0.546) |
| DLST (s) | 0.13 (0.02) | 0.12 (0.02) | 0.169 | -0.134 | (-0.372, 0.115) |
| Stride Time (s) | 1.06 (0.07) | 1.04 (0.06) | **<0.001** | -0.319 | (-0.549, -0.101) |
| Stance Time (s) | 0.66 (0.05) | 0.64 (0.05) | **0.001** | -0.268 | (-0.503, -0.042) |
| Swing Time (s) | 0.41 (0.02) | 0.40 (0.02) | **<0.001** | -0.349 | (-0.587, -0.100) |
| Step Length (cm) | 64.5 (8.2) | 63.5 (8.3) | 0.090 | -0.118 | (-0.290, 0.081) |
| Step Time (s) | 0.53 (0.04) | 0.51 (0.03) | **<0.001** | -0.320 | (-0.551, -0.102) |
| Step Width (cm) | 7.8 (3.0) | 8.2 (3.1) | **0.018** | 0.140 | (-0.025, 0.292) |
| Stride Length (cm) | 127.8 (16.5) | 125.8 (16.7) | 0.066 | -0.123 | (-0.296, 0.078) |
| Walking Speed (cm/s) | 120.4 (19.1) | 121.3 (18.8) | 0.490 | 0.046 | (-0.138, 0.244) |

The variability of step width decreased significantly with DBS treatment *(Table 3)*. In contrast, the variability of DLST, stride time, stance time, and step time all increased significantly, indicating a worsening of most temporal gait variability parameters. The variability of cadence, swing time, step length, and stride length also increased, although these changes were not statistically significantly. The variability of walking speed showed no significant difference with DBS treatment.

**Table 3.** Variability of gait parameters for the Pre-DBS and Post-DBS groups are shown. Statistically significant differences are indicated with bolded numbers.

| Parameters of Gait Variability | Pre-DBS | Post-DBS | p-value (pre-post) | Effect Size (Hedges' g, pre-post) | Effect Size Confidence Intervals |
| --- | --- | --- | --- | --- | --- |
| Cadence (% CV) | 1.64 (0.68) | 1.82 (0.86) | 0.055 | 0.228 | (-0.102, 0.567) |
| DLST (% CV) | 8.29 (2.39) | 9.26 (3.34) | **0.036** | 0.329 | (0.005, 0.644) |
| Stride Time (% CV) | 1.96 (0.61) | 2.15 (0.82) | **0.037** | 0.254 | (-0.069, 0.598) |
| Stance Time (% CV) | 2.70 (0.74) | 3.01 (1.02) | **0.006** | 0.348 | (0.035, 0.690) |
| Swing Time (% CV) | 2.54 (0.68) | 2.78 (1.02) | 0.065 | 0.275 | (-0.025, 0.580) |
| Step Length (% CV) | 3.52 (1.05) | 3.76 (1.31) | 0.085 | 0.200 | (-0.098, 0.478) |
| Step Time (% CV) | 2.44 (0.59) | 2.64 (0.91) | **0.039** | 0.266 | (-0.051, 0.595) |
| Step Width (% CV) | 32.69 (15.30) | 28.30 (12.97) | **0.001** | -0.307 | (-0.551, -0.048) |
| Stride Length (% CV) | 3.11 (1.07) | 3.32 (1.27) | 0.083 | 0.184 | (-0.111, 0.473) |
| Walking Speed (% CV) | 4.00 (2.67) | 3.91 (1.49) | 0.739 | -0.044 | (-0.403, 0.339) |

### 3.3 Relationship between changes in gait performance and location of active contacts

The results of the Kruskal-Wallis test indicate that DBS stimulation location had a greater impact on mean spatial parameters than on mean temporal parameters *(Table 4)*. Specifically, significant effects were observed along the z-axis for mean step length, stride length, and walking speed and along the x-axis for mean walking speed. Mean temporal parameters were not significantly influenced by stimulation location.

**Table 4.**
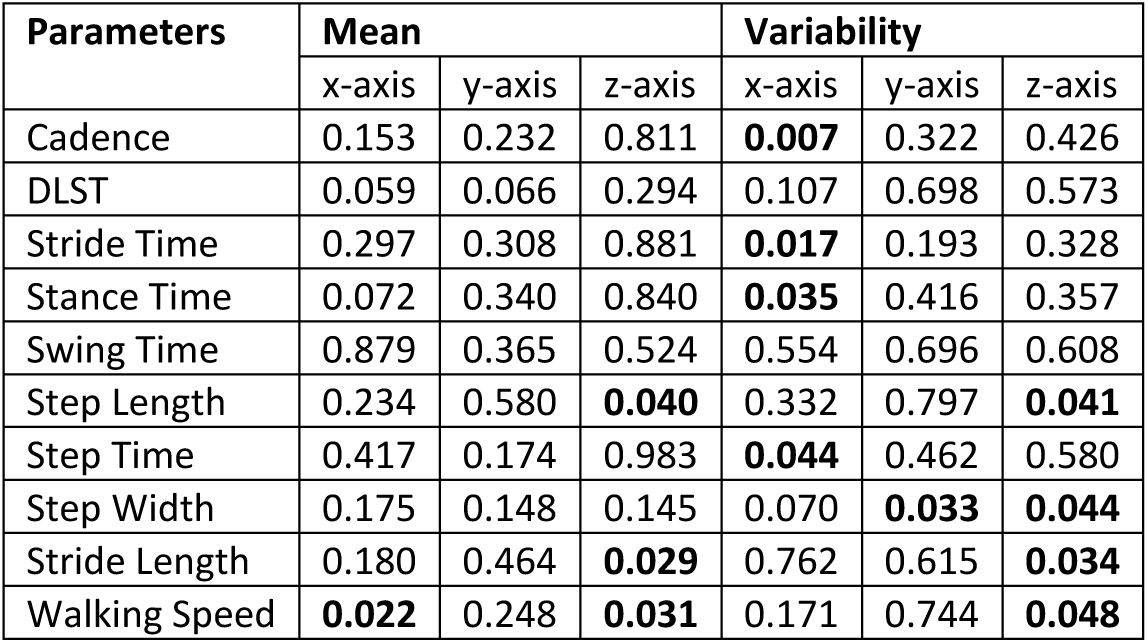
The p-values for the differences in gait parameters according to DBS active contact location along each of the x-, y-, z-axes are provided. Statistically significant differences are indicated with bolded numbers.

Conversely, for gait variability parameters, temporal measures were more influenced by the stimulation location than spatial ones. Specifically, the variabilities of cadence, stride time, step time, and stance time were significantly affected by the location of the active contact along the x-axis. Regarding spatial parameters, variability of step width was influenced by the active contact location along both y- and z-axes, while variability of step length, stride length, and walking speed were influenced along the z-axis.

Within the mean parameter group, the relationship between gait parameters and the location of the active contact exhibited considerable variation across different parameters, with no clear or consistent pattern of parameters showing positive changes (Figure 3, for additional details on the influence of active contact location on each gait parameter, see Supplementary Figure 1). In contrast, within the variability parameters, with the exception of step width, all parameters showed similar associations with the location of active contacts along the x-, y-, and z-axes. Notably, there was a most effective region where the variability of all parameters, except step width and stride length, decreased the most.

**Figure 3.**
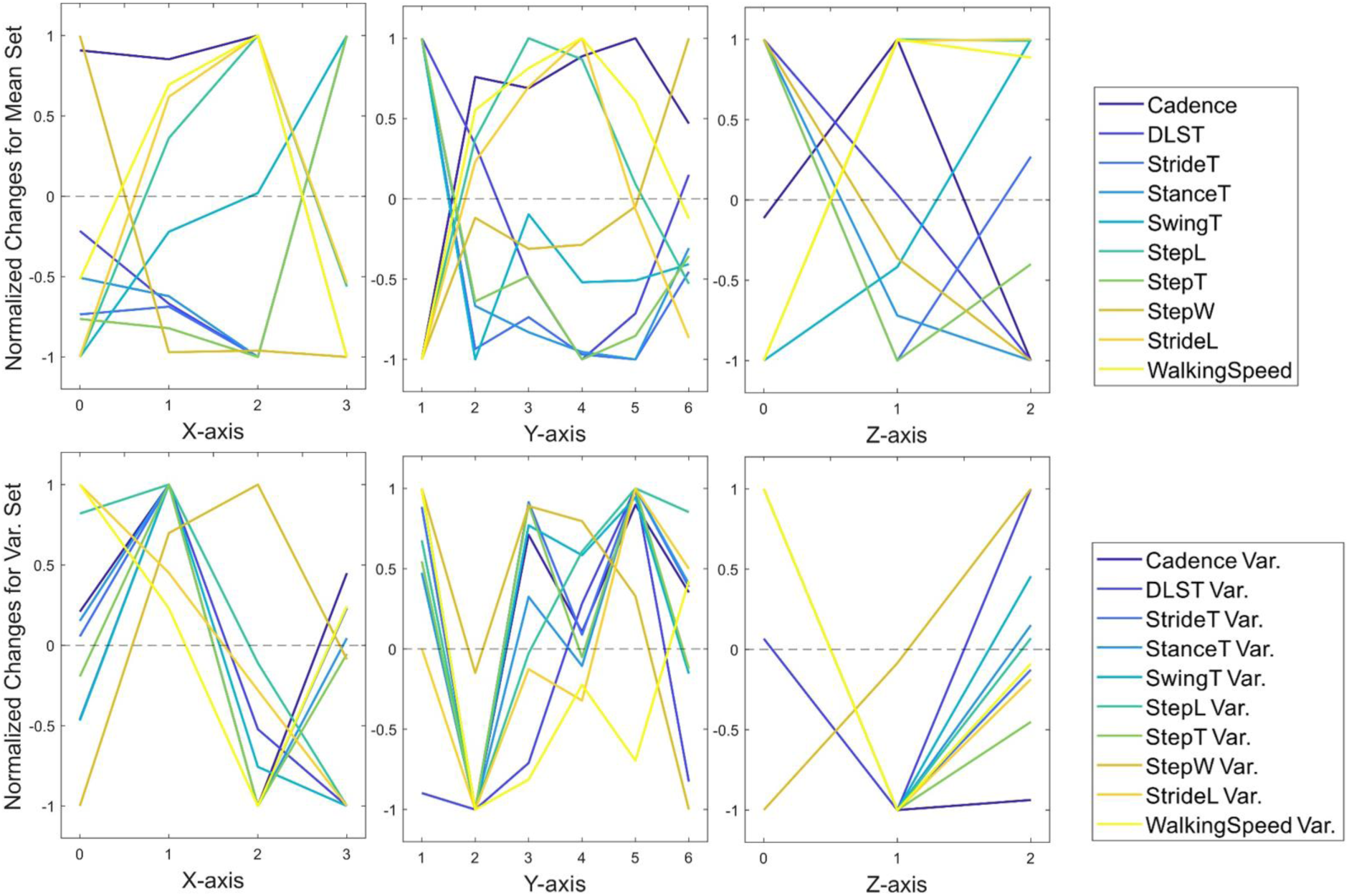
The normalized trends of all gait parameters along the x-, y-, and z-axes for mean (upper) and variability (lower) gait parameters. The normalized trends are the result of normalization between parameters (Post-DBS minus Pre-DBS) to visualize the similarity of responses of different parameters to DBS treatment. The dashed line represents the median change (Post-DBS minus Pre-DBS) among patients.

**Figure 4.**
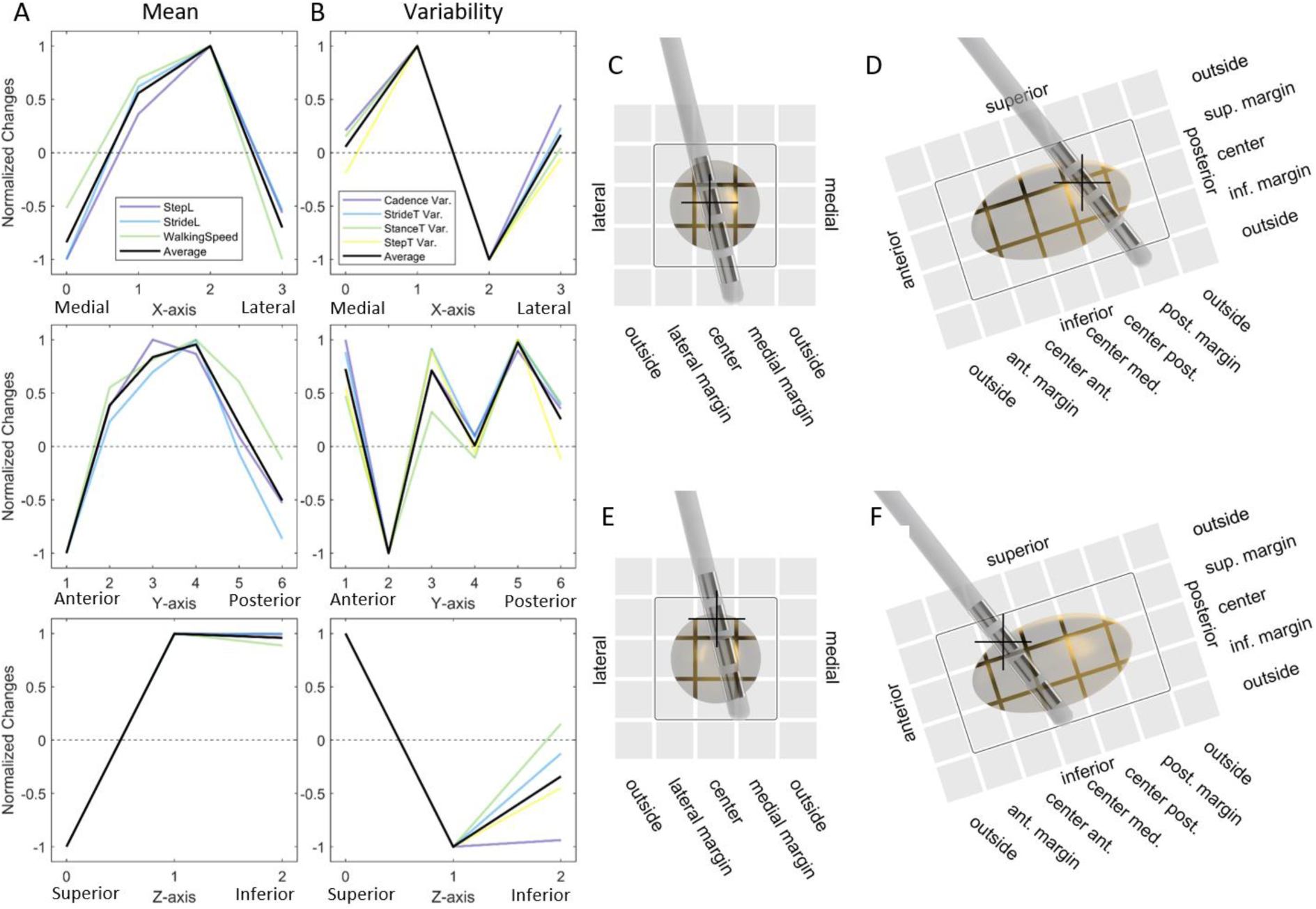
The normalized tendency of the (A) reduced mean set, and (B) reduced variability set, along with the average tendency for each subgroup and direction (black). Schematic of the location of the most effective regions for the (C, D) mean set and (E, F) variability set.

### 3.4 Most effective region for improved gait

The most effective regions for gait performance improvement were analysed separately for the variability and mean parameter groups. Only the parameters significantly affected by stimulation location, according to the Kruskal-Wallis test, were included in the analysis. This resulted in a reduced mean set consisting only of spatial parameters: step length, stride length, and walking speed, and a reduced variability set consisting only of temporal parameters: cadence, stride time, stance time, and step time. Due to its different response to location compared to the other parameters (*c.f.* Figure 3), step width was excluded from both groups.

The most effective region for the reduced mean set was defined as the voxel in which the maximum increase in these parameters occurred, while for the reduced variability set, it was defined as the voxel where the maximum decrease in variability occurred. Both most effective regions were located within the STN but exhibited slight differences in their voxel locations. Therefore, two distinct most effective regions were defined: one for the reduced mean set and one for the reduced variability set (Figure 6C and D). The most effective region for the reduced mean set encompassed voxels (2,4,1), (2,3,1), (2,4,2), and (2,3,2), indicating active electrode contacts within the STN located more posteriorly along the y-axis, superiorly along the z-axis, and near the midline along the x-axis. In contrast, the most effective region for the reduced variability set comprised voxels (2,2,1) and (2,2,2), indicating active electrode contacts within the STN situated more anteriorly along the y-axis, superiorly along the z-axis, and centered along the x-axis (medio-lateral axis).

### 3.5 The difference in stimulation location inside vs. outside the most effective region

The impact of the most effective region on both the mean and variability sets was analysed *(Table 5)*. In the mean set, patients with active contacts located inside the most effective region (inside subgroup), exhibited a greater increase in the means of stride length, step length, and walking speed after DBS treatment compared to those with active contacts located outside the effective region (outside subgroup). Interestingly, the inside subgroup also exhibited a greater decrease in variabilities of stance time, swing time, step length, step time, step width, and walking speed after DBS treatment than the outside subgroup, where the decreases in cadence and stride time were also significant. Moreover, for the inside subgroup, the variability of gait parameters decreased after the DBS surgery, in contrast to the overall trend observed in the entire dataset, where variability generally increased.

**Table 5.**
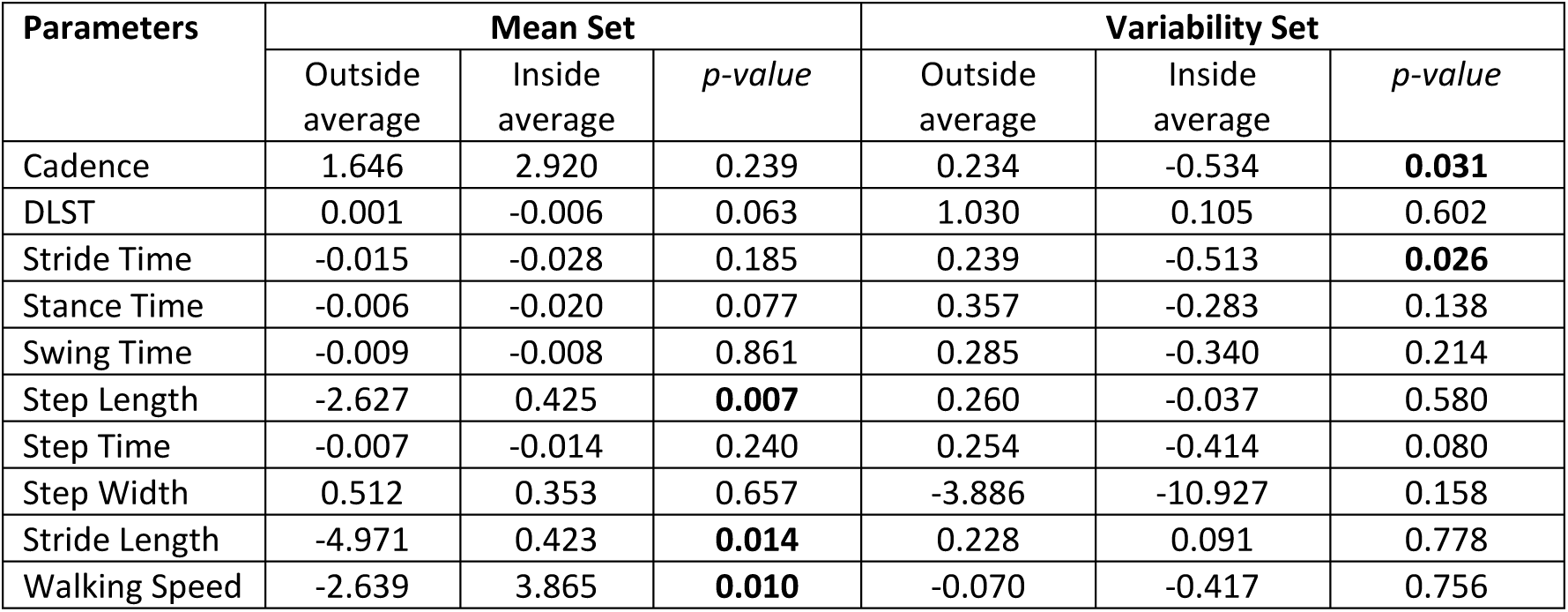
The average of gait parameters changes for patients with active contacts located in the most effective region (inside subgroup) and those without (outside subgroup) after DBS treatment. As long as the p-value for the difference in gait parameters between inside and outside subgroups. Bolded numbers are used to signify statistically significant differences.

| Parameters | Mean Set |  |  | Variability Set |  |  |
| --- | --- | --- | --- | --- | --- | --- |
|  | Outside average | Inside average | *p*-value | Outside average | Inside average | *p*-value |
| Cadence | 1.646 | 2.920 | 0.239 | 0.234 | -0.534 | **0.031** |
| DLST | 0.001 | -0.006 | 0.063 | 1.030 | 0.105 | 0.602 |
| Stride Time | -0.015 | -0.028 | 0.185 | 0.239 | -0.513 | **0.026** |
| Stance Time | -0.006 | -0.020 | 0.077 | 0.357 | -0.283 | 0.138 |
| Swing Time | -0.009 | -0.008 | 0.861 | 0.285 | -0.340 | 0.214 |
| Step Length | -2.627 | 0.425 | **0.007** | 0.260 | -0.037 | 0.580 |
| Step Time | -0.007 | -0.014 | 0.240 | 0.254 | -0.414 | 0.080 |
| Step Width | 0.512 | 0.353 | 0.657 | -3.886 | -10.927 | 0.158 |
| Stride Length | -4.971 | 0.423 | **0.014** | 0.228 | 0.091 | 0.778 |
| Walking Speed | -2.639 | 3.865 | **0.010** | -0.070 | -0.417 | 0.756 |

## 4. Discussion

This study aimed to evaluate the impact of STN-DBS treatment on gait performance in PwPD, to explore the correlations between treatment-related changes and stimulation location within the STN, as well as to define the most effective DBS stimulation regions for maximizing overall gait performance. Our results revealed the following: 1) STN-DBS treatment significantly decreased stride time, stance time, swing time and step time, while significantly increased stepping frequency, but with a compensatory increase in step width, likely to maintain balance. In parallel, STN-DBS decreased the variability of step width but increased the variability of DLST, stride time, stance time, and step time. 2) The location of active stimulation contacts influenced both the magnitude of improvement in mean gait parameters, specifically step length, stride length, and walking speed and the variability of cadence, stride time, stance time, and step width. 3) The most effective region for improving mean step length, stride length, and walking speed was located within the anterior, superior and central (mediolateral direction) region of the STN. The most effective region for improving walking variability was in the posterior, superior and central (mediolateral direction) region of the STN.

### 4.1 The impact of STN-DBS on gait performance

Except for the parameter DLST, the average walking performance, as measured by temporal parameters, changed with STN-DBS. This indicates that STN-DBS treatment can help restore gait function disrupted by PD. However, when considering the effect size of STN-DBS on each parameter, our study revealed values ranging from small (Hedges’ g = 0.2) to medium (Hedges’ g = 0.5), despite being statistically significant according to the t-test. This suggests that the observed effect on each parameter might not be readily perceptible to the human eye. Nevertheless, the effects are not so minimal as to be clinically irrelevant[33]. One potential explanation for the small-to-medium effect sizes could be the superposition of minor alterations across various gait parameters, each contributing to the overall pathological walking pattern.

Our results demonstrated a significant decrease in stance time and swing time, leading to a significant increase in cadence, indicative of faster stepping movements in patients with STN-DBS compared to their pre-treatment status. However, their overall walking speed remained unchanged. These results suggest that PwPD undergoing STN-DBS are capable of executing limb movements more rapidly. However, this increased speed comes at the expense of step length. As a result, patients take quicker, shorter steps and adopt a wider step width, likely as a compensatory strategy to maintain balance, ultimately sustaining a pathological walking pattern. To address this, locomotor and balance training following STN-DBS treatment can be strongly recommended to further enhance walking and balance abilities, supporting patients in adjusting to their new walking patterns, characterized by faster stepping facilitated by STN-DBS.

Extensive evidence supports the relationship between gait variability and walking stability[34, 35]. Elderly individuals typically exhibit higher step width variability compared to younger populations. Furthermore, greater gait variability, whether in spatial or temporal domains, is associated with an increased risk of falls during walking[36, 37]. This relationship is likely due to reduced walking stability and heightened susceptibility to external perturbations when variability is elevated. Conversely, lower variability if often considered an indicator of stable movement. In our study, variability increased across all temporal parameters, as well as in step length and stride length, following STN-DBS treatment. This suggests that PwPD may experience heightened gait variability following DBS surgery, particularly reflecting reduced walking stability in the anteroposterior direction. From a broader perspective, the observed increase in gait variability with STN-DBS appears to indicate a general deterioration in walking performance, despite reductions in step width variability and improvements in other clinical symptoms, such as tremor and rigidity.

### 4.2 The impact of stimulation contact location on gait outcome

This study demonstrates that the influence of stimulation location is particularly pronounced in the mean value of gait parameters stride length, step length, and walking speed, with all of these parameters sharing a comparable region of highest effectiveness. After surgery, the increase in these gait parameters reaches its maximum when the active contact was positioned in the superior, posterior, and central (medio-lateral) region of the STN. The most effective region identified in our study closely aligns with the sweet spot previously identified for optimal alleviation of rigidity and bradykinesia in PwPD (superior, posterior, medial)[23]. The alleviation of rigidity and bradykinesia symptoms may enhance a patient’s ability to initiate and effectively exert movement, thereby improving walking ability[38–40]. This could explain why the most effective region for improving the mean value of spatial gait parameters overlaps with the sweet spot for relieving these clinical symptoms. The sweet spots located at the dorsal interface between the sensorimotor and associative regions of the STN have consistently demonstrated benefits for various motor functions in prior studies[19, 24, 41–45]. Anatomically, the most effective region for improving the mean value of spatial gait parameters appears to correspond to the motor territory of the STN. In the current study, along the superior-inferior axis, stimulating within the STN has been demonstrated to be more effective than stimulation outside the STN, particularly near the thalamus. For the medial-lateral and anterior-posterior planes, the best stimulation location has been suggested to be inside the STN, in close proximity to the substantia nigra.

In the variability set, stimulation location significantly influenced temporal parameters, including cadence, stride time, step time, and stance time. These findings suggest that stimulation of the superior, anterior, and central region within the STN is most effective for decreasing gait variability towards physiological levels. The most effective region for reducing gait variability identified in our study generally aligns with the previously reported sweet spot for improvement of tremor (superior, central in the antero-posterior direction, and lateral STN)[23]. In the elderly, lower gait variability is associated with more stable walking patterns [32]. Previous meta-analyses have also indicated that the dorsolateral STN plays a crucial role in improving rigidity and bradykinesia[22, 41]. Anatomically, the most effective region for reducing gait variability appears to be part of the motor territory of the STN. Based on the present data, stimulation within the STN, rather than outside it near the thalamus, was more effective along the superior-inferior axis. In the medial-lateral and anterior-posterior planes, comparative data from both sides indicate that the most effective stimulation location is adjacent to the globus pallidus, specifically at the connection between the STN and the internal segment of the globus pallidus.

In both the mean and variability sets, the most effective region along the mediolateral axis differs from the findings of previous studies. Our results suggest that PwPD may benefit most from having the central region of the STN stimulated, rather than focusing solely on either the medial or lateral side to address different symptoms. This distinction may, in part, be attributed to the voxel classification approach used in our study. Along the mediolateral axis, the voxels were categorized as: completely outside the STN medially, partially within the STN medially, entirely within the central STN, partially within the STN laterally, and completely outside the STN laterally. As a result, stimulation sites categorized as inside the STN, whether medial, central, or lateral, were all grouped within the voxel classification corresponding to the central STN. In this respect, a higher resolution analysis (i.e. focusing on the precise anatomical location of the active contact) could help elucidate the subtle differences between these regions.

### 4.3 The neurophysiological origin of the most effective stimulation location for gait

Movement generation is a highly complex process that requires the coordination of multiple spinal, bulbar, and suprabulbar structures. Among these, the basal ganglia of the motor circuit play a significant role. Specifically, the STN is a key component of the indirect pathway, transmitting inhibitory signals to the globus pallidus internal segment, thereby exerting inhibition over movement control[1]. Stimulation of the STN can help patients activate and re-calibrate the indirect pathway, improving movement regulation[6]. Hence, the most effective region for managing gait variability, identified on the globus pallidus side of the STN and encompassing the connection between the STN and globus pallidus internal segment, likely corresponds to the anatomical location of the indirect pathway within the STN. Stimulation of this area may enable patients to potentially inhibit redundant movements during walking, thereby fostering a more stable gait pattern with reduced variability. Within the motor circuit, the pars reticulata of the substantia nigra, plays a crucial role in the direct pathway by continuously transmitting inhibitory signals to the thalamus to suppress movement[1]. The substantia nigra-leaning side of the STN, identified as the most effective region in the mean set, likely corresponds to the direct pathway between the pars reticulata and the thalamus. Stimulation of this region may interrupt the direct inhibitory influence of the pars reticulata on the thalamus, facilitating greater thalamic activation and enhancing movement initiation. Consequently, patients may achieve a higher ability to initiate movements, resulting in reduced bradykinesia, longer steps, and increased walking speed.

### 4.4 Limitations

This study has several limitations. First, the patient cohort size may be insufficient for conducting extensive statistical analysis, particularly given the heterogeneity of the recruited sample. Additionally, some voxels included in the anatomical analysis were not represented within our sample, limiting the generalizability of the voxel-based findings. Furthermore, the visually based methodology used to identify the most effective region lacks precision and could benefit from more objective, automated selection methods in future investigations. Finally, the absence of recorded medication intake timings prevented us from accounting for potential medication effects in our analysis.

### 4.5 Conclusions

Overall, from a macroscopic perspective, STN-DBS treatment appears to exacerbate the symptoms of unstable walking. However, in PwPD with active contacts positioned anteriorly and superiorly within the STN, there is evidence of slight improvement in walking performance following DBS treatment. The observed correlation between the location of active contacts for stimulation and gait outcomes suggests that the means of specific spatio-temporal parameters, and the variability of temporal parameters are more responsive to stimulation in specific regions of the STN. Our findings indicate that stimulation within the posterior, superior, and central STN is most effective for enhancing mean spatio-temporal gait parameters, while improvements in gait variability are associated with stimulation in the anterior, superior, and central region of the STN.

This observational, single-center study, conducted under a standardized surgical protocol and approach by the same neurosurgical team, offers key insights into the effects of STN-DBS on gait performance in PwPD from a relatively large patient cohort. The adoption of a voxel-based normalization approach allowed for precise localization of active contacts within the STN, accounting for patient-specific anatomical variability. This method is intuitive for clinicians, enhancing its potential applicability in clinical practice. In future work, we aim to explore advanced modeling approaches, such as Volume of Tissue Activated and Lead-DBS, to further refine the localization of stimulation effects. Additionally, incorporating comparison between generic STN models and each patient’s native STN anatomy could provide further insights into anatomical and functional variability.

In conclusion, integrating detailed pre-operative kinematic assessments alongside standard clinical evaluations for Parkinson’s disease can support the optimisation of stereotactic surgery planning. Targeting active contact locations tailored to each patient’s predominant symptoms and individual pathology holds promise for improving clinical outcomes in PwPD undergoing STN-DBS.

## Abbreviations

PD: Parkinson’s Disease
DBS: Deep Brain Stimulation
STN: Subthalamic Nucleus
PwPD: Patients with Parkinson’s Disease
STN-DBS: Deep brain stimulation of the subthalamic nucleus
UPDRS: the Unified Parkinson Disease Rating Scale
CT: Computed Tomography
MRI: Magnetic Resonance Imaging
AC-PC: Anterior-Commissure to Posterior-Commissure
Pre-DBS: Before DBS Electrode Implantation
Post-DBS: 6 Months after DBS Electrode Implantation
DLST: Double Limb Support Time

## Declarations

### Ethics approval and consent to participate

The study was approved (approval no: 2015-00141) by Zurich Cantonal Ethics Commission and carried out in accordance with the Declaration of Helsinki, the guidelines of Good Clinical Practice, and the Swiss regulatory authority’s requirements. The subjects all provided written, informed consent prior to participation.

### Consent for publication

Consent for publication has been given by all authors.

### Authors’ contributions

DR, ZM, ASH, and LS conceived and designed the study. CB, LS, and MU coordinated the recruitment, pre-surgery, and post-surgery of the participants. DR, NS, and WT coordinated the gait measurements. ASH and LS performed the lead localisation reconstruction. ZM and DR analysed the data. ZM, DR and ASH drafted the manuscript. All the authors reviewed, provided critical opinion, and approved the final version of the manuscript for submission. WRT is the guarantor.

## Data Availability

Due to ethical considerations, open publication of the dataset for further use by researchers outside of the investigating team is not possible. Please contact the corresponding author for collaborations and requests.

## References

1. Poewe, W., et al., Parkinson disease. Nat Rev Dis Primers, 2017. 3: p. 17013.

2. Halliday, G.M. and H. McCann, The progression of pathology in Parkinson’s disease. Ann N Y Acad Sci, 2010. 1184: p. 188–95.

3. Bratsos, S., D. Karponis, and S.N. Saleh, Efficacy and Safety of Deep Brain Stimulation in the Treatment of Parkinson’s Disease: A Systematic Review and Meta-analysis of Randomized Controlled Trials. Cureus, 2018. 10(10): p. e3474.

4. Ravi, D.K., et al., Does Subthalamic Deep Brain Stimulation Impact Asymmetry and Dyscoordination of Gait in Parkinson’s Disease? Neurorehabil Neural Repair, 2021. 35(11): p. 1020–1029.

5. Deuschl, G. and Y. Agid, Subthalamic neurostimulation for Parkinson’s disease with early fluctuations: balancing the risks and benefits. Lancet Neurol, 2013. 12(10): p. 1025–34.

6. Mahlknecht, P., et al., How Does Deep Brain Stimulation Change the Course of Parkinson’s Disease? Mov Disord, 2022. 37(8): p. 1581–1592.

7. Wilkins, K.B., et al., Beta Burst-Driven Adaptive Deep Brain Stimulation Improves Gait Impairment and Freezing of Gait in Parkinson’s Disease. medRxiv, 2024.

8. Collomb-Clerc, A. and M.L. Welter, Effects of deep brain stimulation on balance and gait in patients with Parkinson’s disease: A systematic neurophysiological review. Neurophysiol Clin, 2015. 45(4-5): p. 371–88.

9. Cossu, G. and M. Pau, Subthalamic nucleus stimulation and gait in Parkinson’s Disease: a not always fruitful relationship. Gait Posture, 2017. 52: p. 205–210.

10. Huhn, M., et al., Comparison of the Long-Term Efficacy of Targeting the Subthalamic Nucleus Versus the Globus Pallidus Interna for Deep Brain Stimulation Treatment of Motor Dysfunction in Patients With Parkinson’s Disease: A Meta-Analysis Study. Parkinsons Dis, 2024. 2024: p. 5157873.

11. Azgomi, H.F., et al., Modeling and Optimizing Deep Brain Stimulation to Enhance Gait in Parkinson’s Disease: Personalized Treatment with Neurophysiological Insights. medRxiv, 2024.

12. Navratilova, D., et al., Deep Brain Stimulation Effects on Gait Pattern in Advanced Parkinson’s Disease Patients. Front Neurosci, 2020. 14: p. 814.

13. Kroneberg, D., et al., Kinematic Effects of Combined Subthalamic and Dorsolateral Nigral Deep Brain Stimulation in Parkinson’s Disease. J Parkinsons Dis, 2024. 14(2): p. 269–282.

14. Werner, L.M., A. Schnitzler, and J. Hirschmann, Subthalamic Nucleus Deep Brain Stimulation in the Beta Frequency Range Boosts Cortical Beta Oscillations and Slows Down Movement. J Neurosci, 2025. 45(9).

15. Su, Z.H., et al., Deep Brain Stimulation and Levodopa Affect Gait Variability in Parkinson Disease Differently. Neuromodulation, 2023. 26(2): p. 382–393.

16. Horn, A., et al., Lead-DBS v2: Towards a comprehensive pipeline for deep brain stimulation imaging. Neuroimage, 2019. 184: p. 293–316.

17. Caire, F., et al., A systematic review of studies on anatomical position of electrode contacts used for chronic subthalamic stimulation in Parkinson’s disease. Acta Neurochir (Wien), 2013. 155(9): p. 1647–54; discussion 1654.

18. Zhang, F., et al., Relationship between electrode position of deep brain stimulation and motor symptoms of Parkinson’s disease. BMC Neurol, 2021. 21(1): p. 122.

19. Haegelen, C., et al., Functional atlases for analysis of motor and neuropsychological outcomes after medial globus pallidus and subthalamic stimulation. PLoS One, 2018. 13(7): p. e0200262.

20. Dembek, T.A., et al., Probabilistic sweet spots predict motor outcome for deep brain stimulation in Parkinson disease. Ann Neurol, 2019. 86(4): p. 527–538.

21. Reich, M., et al., Probabilistic mapping of gait changes after STNDBS for Parkinson’s disease. 2022.

22. Boutet, A., et al., Sign-specific stimulation ‘hot’ and ‘cold’ spots in Parkinson’s disease validated with machine learning. Brain Commun, 2021. 3(2): p. fcab027.

23. Akram, H., et al., Subthalamic deep brain stimulation sweet spots and hyperdirect cortical connectivity in Parkinson’s disease. Neuroimage, 2017. 158: p. 332–345.

24. Malaga, K.A., et al., Atlas-independent, N-of-1 tissue activation modeling to map optimal regions of subthalamic deep brain stimulation for Parkinson disease. Neuroimage Clin, 2021. 29: p. 102518.

25. Hilliard, J.D., R.C. Frysinger, and W.J. Elias, Effective subthalamic nucleus deep brain stimulation sites may differ for tremor, bradykinesia and gait disturbances in Parkinson’s disease. Stereotact Funct Neurosurg, 2011. 89(6): p. 357–64.

26. König, N., et al., Is gait variability reliable? An assessment of spatio-temporal parameters of gait variability during continuous overground walking. Gait & Posture, 2014. 39(1): p. 615–617.

27. Efron, B.a.R.T., An Introduction to the Bootstrap. 1994.

28. Ho, J., et al., Moving beyond P values: data analysis with estimation graphics. Nat Methods, 2019. 16(7): p. 565–566.

29. Enzmann, D., Notes on Effect Size Measures for the Difference of Means From Two Independent Groups: The Case of Cohen’s d and Hedges’ g. 2015.

30. Mirelman, A., et al., Gait impairments in Parkinson’s disease. Lancet Neurol, 2019. 18(7): p. 697–708.

31. Iansek, M.E.M.H.M.D., The biomechanics and motor control of gait in Parkinson disease. 2001.

32. Konig, N., et al., Revealing the quality of movement: A meta-analysis review to quantify the thresholds to pathological variability during standing and walking. Neurosci Biobehav Rev, 2016. 68: p. 111–119.

33. Coe, R., It’s the Effect Size, Stupid What effect size is and why it is important. 2012.

34. Skiadopoulos, A. and N. Stergiou, Risk-of-falling related outcomes improved in community-dwelling older adults after a 6-week sideways walking intervention: a feasibility and pilot study. BMC Geriatr, 2021. 21(1): p. 60.

35. Yang, F. and Y.C. Pai, Can stability really predict an impending slip-related fall among older adults? Journal of Biomechanics, 2014. 47(16): p. 3876–3881.

36. Hausdorff, J.M., et al., Increased gait unsteadiness in community-dwelling elderly fallers. Archives of Physical Medicine and Rehabilitation, 1997. 78(3): p. 278–283.

37. Hausdorff, J.M., D.A. Rios, and H.K. Edelberg, Gait variability and fall risk in community-living older adults: A 1-year prospective study. Archives of Physical Medicine and Rehabilitation, 2001. 82(8): p. 1050–1056.

38. Buurke, T.J.W., et al., The effect of walking with reduced trunk motion on dynamic stability in healthy adults. Gait Posture, 2023. 103: p. 113–118.

39. Dotov, D., et al., Coordination Rigidity in the Gait, Posture, and Speech of Persons with Parkinson’s Disease. J Mot Behav, 2023. 55(4): p. 394–409.

40. Onder, H., et al., The gait parameters in patients with Parkinson’s Disease under STN-DBS therapy and associated clinical features. Neurol Res, 2023. 45(8): p. 779–785.

41. Butson, C.R., et al., Probabilistic analysis of activation volumes generated during deep brain stimulation. Neuroimage, 2011. 54(3): p. 2096–104.

42. Conrad, E.C., et al., Atlas-Independent, Electrophysiological Mapping of the Optimal Locus of Subthalamic Deep Brain Stimulation for the Motor Symptoms of Parkinson Disease. Stereotact Funct Neurosurg, 2018. 96(2): p. 91–99.

43. Herzog, J., et al., Most effective stimulation site in subthalamic deep brain stimulation for Parkinson’s disease. Mov Disord, 2004. 19(9): p. 1050–4.

44. Maks, C.B., et al., Deep brain stimulation activation volumes and their association with neurophysiological mapping and therapeutic outcomes. J Neurol Neurosurg Psychiatry, 2009. 80(6): p. 659–66.

45. Yokoyama, T., et al., Relationship of stimulation site location within the subthalamic nucleus region to clinical effects on parkinsonian symptoms. Stereotact Funct Neurosurg, 2006. 84(4): p. 170–5.

